# Post-COVID-19 syndrome and related dysautonomia affect patients’ life and work productivity

**DOI:** 10.1101/2023.03.15.23287298

**Authors:** Petros Galanis, Aglaia Katsiroumpa, Irene Vraka, Katerina Kosiara, Olga Siskou, Olympia Konstantakopoulou, Theodoros Katsoulas, Parisis Gallos, Daphne Kaitelidou

## Abstract

**Background:** A significant percentage of COVID-19 patients experience post-COVID-19 symptoms and signs. Post-COVID-19 syndrome affects physical and mental health of patients in several ways.

**Aim:** To investigate the impact of post-COVID-19 syndrome and related dysautonomia on patients’ life and work productivity.

**Methods:** We conducted a cross-sectional study in Greece using an online questionnaire. Study population included 108 workers over 18 years old that have been diagnosed with post-COVID-19 syndrome. Patients were recruited from the Long COVID Greece patients’ society. We measured demographic and clinical characteristics of patients, resilience, and social support.

**Results:** Among patients, 68.5% stated that post-COVID-19 syndrome affected their daily life to a great extent, 25% to a moderate level, and 6.5% to a small extent. Moreover, 56.5% stated that post-COVID-19 syndrome affected their work productivity to a great extent, 27.8% to a moderate level, and 15.7% to a small extent. Multivariable analysis identified that females and patients with post-COVID-19 dysautonomia had more problems in their daily life. Moreover, increased duration of COVID-19 symptoms was associated with increased daily problems. Increased resilience was related with fewer problems in daily life. Also, we found that patients with post-COVID-19 dysautonomia had less work productivity. Moreover, increased duration of COVID-19 symptoms was associated with more problems in work. Resilience was related with increased work productivity.

**Conclusions:** Post-COVID-19 syndrome and related dysautonomia affect significantly patients’ daily and work life. Also, resilience is an important preventive factor improving patients’ life. Policy makers should develop and implement educational programs to improve patients’ life. Healthcare professionals should be aware of the post-COVID-19 syndrome and its consequences in order to understand post-COVID-19 patients and their problems.

## Introduction

A significant percentage of COVID-19 patients (30%) and hospitalized COVID-19 patients (80%) experience post-COVID-19 symptoms and signs (Huang et al., 2021; Tenforde et al., 2020). The World Health Organization defines post-COVID-19 syndrome as the condition where patients experience symptoms and signs more than 12 weeks after SARS-CoV-2 infection without an alternative diagnosis to be possible (World Health Organization, 2023). A variety of neurological and neuropsychiatric post-COVID-19 symptoms have been established in the literature such as fatigue, brain fog, memory issues, cognitive impairment, headache, sleep disturbances, attention disorder etc. (Ceban et al., 2022; Chadda et al., 2022; Premraj et al., 2022). Moreover, post-COVID-19-related dysautonomia increase physical complaints among patients causing fatigue, heart rate variability, and orthostatic hypotension (Barizien et al., 2021; Dani et al., 2021; Miglis et al., 2020).

Post-COVID-19 syndrome affects also mental health of patients. In particular, several mental health issues are common among post-COVID-19 patients such as anxiety, depression, and post-traumatic stress disorder (European Centre for Disease Prevention and Control, 2022; Shanbehzadeh et al., 2021). Moreover, literature suggests that post-COVID-19 patients experience a poor quality of life since they have problems regarding usual activities, mobility, and self-care (Malik et al., 2022; Premraj et al., 2022). There are great differences in frequency of mental health issues among studies worldwide due to different study designs and definitions (Shanbehzadeh et al., 2021).

Since the post-COVID-19 syndrome affects patients’ life in several ways we investigated the impact of post-COVID-19 syndrome and related dysautonomia on patients’ life and work productivity.

## Methods

### Study design

Study design and methods were described in detail elsewhere (Galanis et al., 2023a, 2023b). In brief, we conducted a cross-sectional study in Greece using an online questionnaire between November 2022 and January 2023. Study population included 108 workers over 18 years old that have been diagnosed with post-COVID-19 syndrome. Patients were recruited from the Long COVID Greece patients’ society (Long COVID Greece, 2023). Our participants were working before and after the post-COVID-19 syndrome in order to assess the impact of disease on their productivity.

We measured the following demographic and clinical characteristics of patients: gender, age, chronic disease, post-COVID-19 dysautonomia, duration of COVID-19 symptoms, hospitalization in COVID-19 ward and COVID-19 intensive care unit, and anxiety disorders and depression before the post-COVID-19 syndrome.

We used the Brief Resilience Scale (BRS) to measure patients’ resilience (Smith et al., 2008). Also, we used the Multidimensional Scale of Perceived Social Support (MSPSS) to measure social support that patients receive (Zimet et al., 1988).

As study outcomes we measured the effect of post-COVID-19 syndrome on patients’ daily life and work productivity. We measured these two outcomes in a scale from 0 (not at all problems) to 10 (extreme problems).

### Ethical issues

We conducted our study applying the guidelines of the Declaration of Helsinki. Also, our study protocol was approved by the Ethics Committee of Faculty of Nursing, National and Kapodistrian University of Athens (reference number; 420, 10 October 2022). Moreover, participants gave their informed consent to participate in our study.

### Statistical analysis

We use mean and standard deviation to present continuous variables. Also, we use numbers and percentages to describe categorical variables. Demographic and clinical characteristics of patients, resilience, and social support were the independent variables. We considered the effect of post-COVID-19 syndrome on patients’ daily life and work productivity as the outcome variables. Since the outcome variables followed the normal distribution, we constructed univariate and multivariable linear regression models. We present the unadjusted and adjusted coefficients beta, 95% confidence intervals (CI) and p-values. P-values less than 0.05 were considered as statistically significant. We performed the statistical analysis with the IBM SPSS 21.0 (IBM Corp. Released 2012. IBM SPSS Statistics for Windows, Version 21.0. Armonk, NY: IBM Corp.).

## Results

Demographic and clinical characteristics of patients with post-COVID-19 syndrome are shown in Table 1. Mean age of patients was 43.5 years. Most of patients were females (76.9%). Among patients, 28.7% suffered from a chronic disease, 15.7% had anxiety disorders before post-COVID-19 syndrome, and 10.2% had depression before the syndrome. Regarding COVID-19, almost half of the patients had post-COVID-19-related dysautonomia (48.1%), 14.8% have been hospitalized in COVID-19 ward, and 2.8% have been hospitalized in COVID-19 intensive care unit. Mean duration of COVID-19 symptoms was 11.8 months.

**Table 1.**
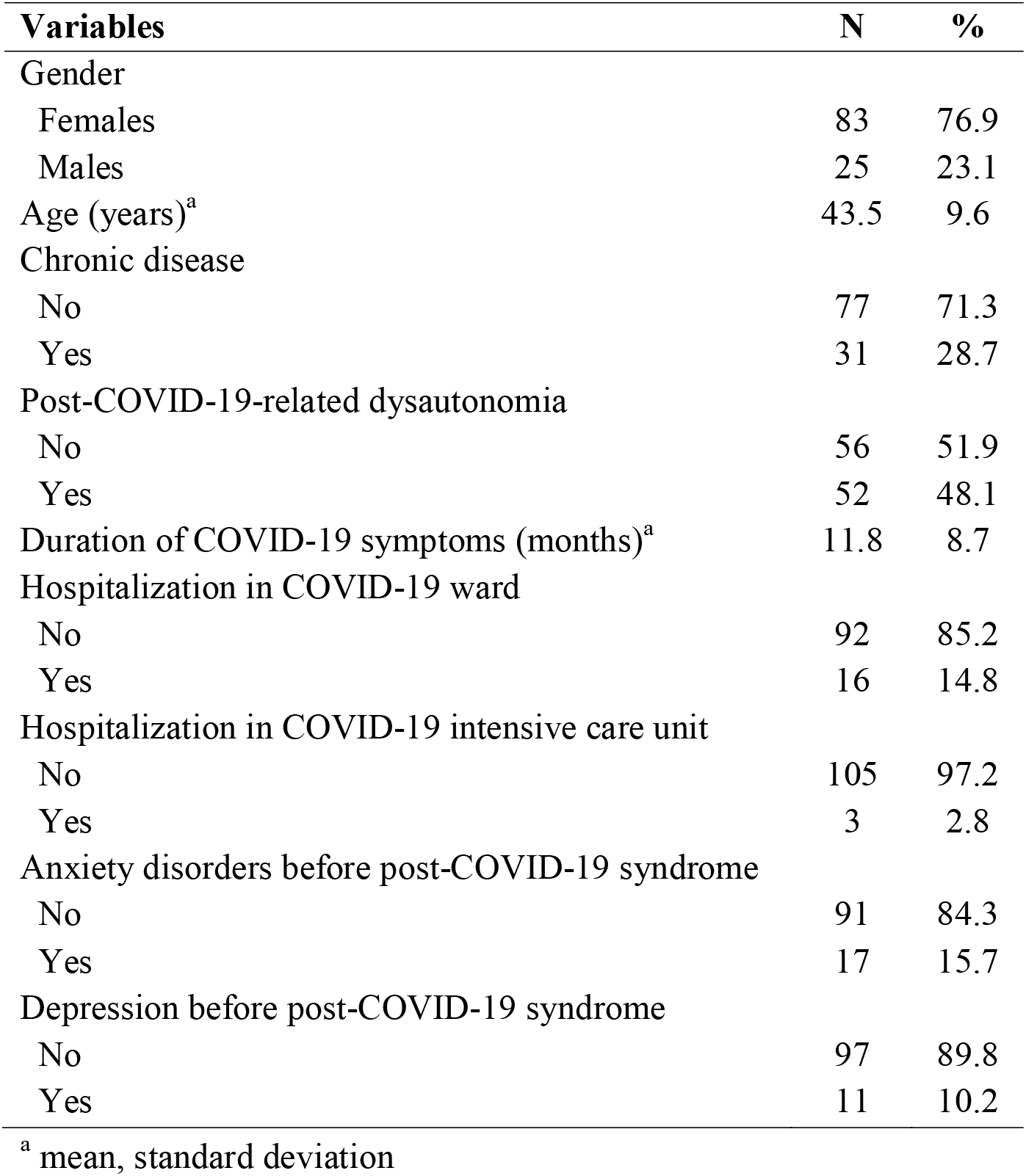
Demographic and clinical characteristics of patients with post-COVID-19 syndrome (N=108).

Mean resilience score was 3.32 (standard deviation = 0.86, median = 3.33) indicating a moderate level of patients’ resilience. Also, mean social support score was 5.36 (standard deviation = 1.44, median = 5.67) indicating a moderate level of social support.

Mean effect score of post-COVID-19 syndrome on patients’ daily life was 7.16 (standard deviation = 2.49, median = 8). Among patients, 68.5% (n=74) stated that post-COVID-19 syndrome affected their daily life to a great extent, 25% (n=27) to a moderate level, and 6.5% (n=7) to a small extent.

Mean effect score of post-COVID-19 syndrome on patients’ work productivity was 6.18 (standard deviation = 2.61, median = 7). Among patients, 56.5% (n=61) stated that post-COVID-19 syndrome affected their work productivity to a great extent, 27.8% (n=30) to a moderate level, and 15.7% (n=17) to a small extent.

Relationship between independent variables and patients’ daily life after the post-COVID-19 syndrome is shown in Table 2. Multivariable analysis identified that females (coefficient beta=1.23, 95% CI=0.19 to 2.27, p-value=0.021) and patients with post-COVID-19 dysautonomia (coefficient beta=0.96, 95% CI=0.06 to 1.85, p-value=0.036) had more problems in their daily life. Moreover, increased duration of COVID-19 symptoms was associated with increased daily problems (coefficient beta=0.07, 95% CI=0.01 to 0.12, p-value=0.014). Resilience was a protective factor, since increased resilience was related with less problems in daily life (coefficient beta=-0.56, 95% CI=-1.06 to -0.05, p-value=0.031).

**Table 2.** Univariate and multivariable linear regression analysis with patients’ daily life after the post-COVID-19 syndrome as the dependent variable.

| Independent variables | Univariate model |  | Multivariable model |  |
| --- | --- | --- | --- | --- |
|  | Unadjusted coefficient beta<br>(95% CI) | P-value | Adjusted coefficient beta<br>(95% CI)<sup>a</sup> | P-value |
| Gender (females vs. males) | 1.66 (0.57 to 2.75) | 0.003 | 1.23 (0.19 to 2.27) | 0.021 |
| Age (years) | -0.02 (-0.07 to 0.03) | 0.396 | -0.008 (-0.06 to 0.04) | 0.752 |
| Chronic disease (yes vs. no) | 0.22 (-0.84 to 1.28) | 0.680 | 0.54 (-0.49 to 1.57) | 0.300 |
| Post-COVID-19 dysautonomia (yes vs. no) | 1.44 (0.52 to 2.36) | 0.002 | 0.96 (0.06 to 1.85) | 0.036 |
| Duration of COVID-19 symptoms | 0.09 (0.03 to 0.14) | 0.001 | 0.07 (0.01 to 0.12) | 0.014 |
| Hospitalization in COVID-19 ward (yes vs. no) | 0.19 (-1.16 to 1.53) | 0.786 | 0.71 (-0.65 to 2.06) | 0.304 |
| Anxiety disorders before post-COVID-19 syndrome (yes vs. no) | 1.14 (-0.16 to 2.44) | 0.084 | 0.24 (-1.25 to 1.74) | 0.748 |
| Depression before post-COVID-19 syndrome (yes vs. no) | 1.24 (-0.32 to 2.81) | 0.119 | 0.70 (-1.05 to 2.44) | 0.430 |
| Resilience | -0.58 (-1.13 to -0.02) | 0.041 | -0.56 (-1.06 to -0.05) | 0.031 |
| Social support | -0.25 (-0.58 to 0.09) | 0.143 | -0.13 (-0.47 to 0.21) | 0.444 |

CI: confidence interval; <sup>a</sup> p-value for ANOVA<0.001; R<sup>2</sup> for the final multivariable model was 19.1%

Linear regression analysis with patients’ work productivity after the post-COVID-19 syndrome as the dependent variable is shown in Table 3. We found that patients with post-COVID-19 dysautonomia (coefficient beta=1.21, 95% CI=0.24 to 2.18, p-value=0.015) had less work productivity. Moreover, increased duration of COVID-19 symptoms was associated with more problems in work (coefficient beta=0.06, 95% CI=0.001 to 0.11, p-value=0.049). Resilience was related with increased work productivity (coefficient beta=-0.57, 95% CI=-1.12 to -0.02, p-value=0.042).

**Table 3.** Univariate and multivariable linear regression analysis with patients’ work productivity after the post-COVID-19 syndrome as the dependent variable.

| Independent variables | Univariate model |  | Multivariable model |  |
| --- | --- | --- | --- | --- |
|  | Unadjusted coefficient beta<br>(95% CI) | P-value | Adjusted coefficient beta<br>(95% CI)<sup>a</sup> | P-value |
| Gender (females vs. males) | 0.39 (-0.80 to 1.57) | 0.52 | 0.14 (-1.05 to 1.32) | 0.818 |
| Age (years) | -0.02 (-0.07 to 0.04) | 0.52 | -0.004 (-0.06 to 0.05) | 0.888 |
| Chronic disease (yes vs. no) | 0.65 (-0.44 to 1.75) | 0.24 | 0.88 (-0.25 to 1.99) | 0.125 |
| Post-COVID-19 dysautonomia (yes vs. no) | 1.52 (0.56 to 2.47) | 0.002 | 1.21 (0.24 to 2.18) | 0.015 |
| Duration of COVID-19 symptoms | 0.07 (0.01 to 0.13) | 0.014 | 0.06 (0.001 to 0.11) | 0.049 |
| Hospitalization in COVID-19 ward (yes vs. no) | 0.16 (-1.25 to 1.57) | 0.822 | 0.48 (-0.99 to 1.96) | 0.517 |
| Anxiety disorders before post-COVID-19 syndrome (yes vs. no) | 1.19 (-0.17 to 2.54) | 0.085 | 0.59 (-1.04 to 2.22) | 0.473 |
| Depression before post-COVID-19 syndrome (yes vs. no) | 0.92 (-0.73 to 2.56) | 0.271 | 0.49 (-1.41 to 2.39) | 0.610 |
| Resilience | -0.58 (-1.16 to -0.003) | 0.049 | -0.57 (-1.12 to -0.02) | 0.042 |
| Social support | -0.22 (-0.57 to 0.13) | 0.209 | -0.07 (-0.44 to 0.29) | 0.695 |

CI: confidence interval; <sup>a</sup> p-value for ANOVA<0.001; R<sup>2</sup> for the final multivariable model was 12.3%

## Discussion

We conducted a cross-sectional study in Greece to investigate the impact of post-COVID-19 syndrome and related dysautonomia on patients’ life and work productivity. Moreover, we examined several demographic and clinical characteristics of post-COVID-19 patients, resilience, and social support as possible determinants of daily life and work productivity.

Our findings showed that a high percentage of post-COVID-19 patients experienced significant problems in their daily life (68.5%) and their work (56.5%). Several studies confirm this finding since quality of life among post-COVID-19 patients drops significantly after the syndrome (Garrigues et al., 2020; Huang et al., 2021; Tabacof et al., 2022; Taboada et al., 2021). Since post-COVID-19 syndrome is a new medical condition without therapy and patients have to deal with unknown consequences it is reasonable that they experience negative feelings.

Moreover, we found that post-COVID-19-related dysautonomia increased patients’ problems. Literature suggests that levels of fatigue is higher among patients with post-COVID-19 dysautonomia (Barizien et al., 2021; Chadda et al., 2022; Lo, 2021; Malik et al., 2022). Also, post-COVID-19 patients with dysautonomia showed reduced work rate and exercise (Ladlow et al., 2022). The major clinical manifestation of post-COVID-19 dysautonomia is fatigue and therefore patients show limited capability to work and exercise. The frequent development of brain fog among post-COVID-19 patients is also a factor that affects their ability to work and adapt to daily activities (Azcue et al., 2022; Orfei et al., 2022). Impaired memory or multitasking that occur frequent among post-COVID-19 patients with brain fog are risk factors for a poorer quality of life at work (Chatys-Bogacka et al., 2022).

Our study showed that there was a negative relationship between resilience and daily life and work productivity. In particular, patients with lower levels of resilience had more problems in their daily life and work. Several meta-analyses confirm the positive effect of resilience on individuals’ life. For example, higher levels of resilience decrease psychological distress in COVID-19 patients (Jeamjitvibool et al., 2022). Moreover, during the pandemic resilience acted as a protective factor maintaining good quality of life among people (Aldhahi et al., 2021; Javellana et al., 2022; Koivunen et al., 2022) Also, there is a negative relationship between resilience and anxiety and depression among patients with a somatic illness or health problem (Färber & Rosendahl, 2018). A meta-analysis found that resilience improved positive indicators of mental health and decreased negative indicators of mental health in the general population (Hu et al., 2015).

We found that increased duration of COVID-19 symptoms was associated with increased daily problems and less work productivity. Symptom duration, hospital admission, and length of hospitalization are associated with increased severity of COVID-19 (Tan et al., 2022). Also, among demographic characteristics we found that females had more problems in their daily life. Literature confirms this finding since females COVID-19 patients had worse quality of life than males during the pandemic (Levy, 2022; Zwar et al., 2023). Moreover, in general, prevalence of poor quality of life, disability, anxiety, and depression is higher among female patients than males (Crispino et al., 2020; Heller et al., 2014).

### Limitations

Our study had several limitations. First, we conducted a cross-sectional study and therefore we cannot establish causal relationships between independent variables and patients’ life and work productivity. Second, we used a self-completed questionnaire to assess patients’ attitudes. Thus, an information bias could be emerged in our study. Third, we measured patients’ attitudes in a particular time. It is probable that these attitudes could change over time. Thus, longitudinal studies should be conducted in order to get more valid results. Fourth, our convenience sample was limited since it is difficult to approach post-COVID-19 patients and participate in studies. Further studies with bigger and more representative samples could add significant information. Fifth, we measured the effect of several demographic and clinical characteristics but other factors could also affect patients’ life and work productivity.

## Conclusions

In conclusion, we found that post-COVID-19 syndrome and related dysautonomia affect significantly patients’ daily and work life. Also, our study showed that resilience was an important preventive factor improving patients’ life. Since post-COVID-19 syndrome is prevalent among COVID-19 patients, policy makers should develop and implement educational programs to improve patients’ life. Also, special attention should be given to the work issues of post-COVID-19 patients because their ability to work is decreased. For example, teleworking could help post-COVID-19 patients to continue working in a productive way. In that way, patients will feel useful and productive improving their quality of life. Healthcare professionals should be aware of the post-COVID-19 syndrome and its consequences in order to understand post-COVID-19 patients and their problems. Therefore, healthcare workers should help post-COVID-19 patients to improve their life by adjusting in a new and difficulty situation such as the post-COVID-19 syndrome.

## Data Availability

All data produced in the present study are available upon reasonable request to the authors

## Acknowledgments

The authors would like to thank the clinicians and Long COVID Greece society members who contributed their time, thoughts, and experiences to this
study.

